# Heat treatment for reuse of disposable respirators during Covid-19 pandemic: Is filtration and fit adversely affected?

**DOI:** 10.1101/2020.04.22.20074989

**Authors:** Miranda Loh, Ross Clark, John Cherrie

**Affiliations:** Institute of Occupational Medicine, Edinburgh, United Kingdom; Institute of Biological Chemistry, Biophysics and Bioengineering, Heriot Watt University, Riccarton, Edinburgh EH14 4AS, UK

**Keywords:** Respirator, Covid-19, filtering face piece, quantitative fit test, filtration efficiency, total inward leakage, personal protective equipment, respiratory protective equipment

## Abstract

A number of methods for decontaminating disposable filtering face piece respirators have been explored for use in health care settings during epidemics where respirators are in short supply, such as the current Covid-19 pandemic. Heating to temperatures above 65°C has been shown to successfully inactivate the SARS-CoV-2 virus on various surfaces. Ovens or similar heating devices are likely already widely available in hospitals globally. We did a quantitative fit test on nine models of FFP2 and FFP3 respirators before and after heat treatment in an oven. These included both flat fold and moulded cup styles. All passed the initial fit test, and all but two passed the post-treatment fit test. This study demonstrates that FFP respirators can still retain both filtration efficiency and fit after wear and heat treatment, but that it is necessary to understand the probability for failure of fit after decontamination. Heat shows promise as a simple and effective way of treating FFP respirators. Further evaluation of longer-term wear and disinfection effectiveness of heat treatment should be done before widespread use.

## Introduction

The current Covid-19 pandemic is not only a public health crisis but also an occupational health crisis; numerous countries face shortages of personal protective equipment (PPE). In healthcare settings, disposable filtering face piece (FFP) respirators that have filtration efficiencies of 95% or 99% (N95 in USA and FFP3 in Europe, respectively) are needed for frontline workers treating patients with Covid-19. The SARS-CoV-2 virus responsible for the disease can be shed and transmitted through the respiratory route, especially in close contact, and therefore any shortage of respiratory protective equipment (RPE) is detrimental to the health of frontline workers.

In the past, studies have explored methods for decontaminating and reusing FFP respirators, but this has not been put into common practice (Heimbuch et al., 2011; Lindsley et al., 2015; Lore et al., 2012; Viscusi et al., 2009). Now, numerous initiatives (“N95DECON - A scientific consortium for data-driven study of N95 FFR decontamination,” 2020) are underway to determine the effectiveness of decontamination methods in 1) disinfecting respirators and 2) maintaining both the filtration efficiency and fit of the respirator. Respirators that meet filtration standards (N95 or FFP3) may still not confer adequate protection due to inward leakage of a respirator poorly fitted to the wearer’s face (Cherrie et al., 2018). Even fit-tested respirators may lose adequate fit after being worn, thus disposable FFP respirators are normally meant for single use only. Retaining fit is as important to maintaining RPE effectiveness as filtration for any decontamination method. Decontamination methods that have been tested and found able to maintain FFP effectiveness without leaving a residue that may be either a nuisance or hazardous to the wearer include UV germicidal irradiation (UVGI), hydrogen peroxide vapour, and heat (CDC, 2020; “Decontamination Methods for 3M N95 Respirators,” 2020; Lindsley et al., 2015; Viscusi et al., 2009).

While effective, methods such as UVGI and hydrogen peroxide vapour may not be readily available in a care facility. The SARS-CoV-2 virus has been shown to be rendered inactive at temperatures of 70°C (Chin et al., 2020). Heat is a relatively easy and accessible method of decontamination. Filtration efficiency up to 110°C dry heat was still retained for three unused N95 models in one experiment (Viscusi et al., 2009). However there is no information on how this affects total inward leakage (which accounts for both filtration and fit) may be affected by heat. We tested a small set of FFP respirators donated by hospitals and various other organisations to evaluate whether heat treatment would adversely affect the ability of the respirator to pass a quantitative fit test after one wear and subsequent heat treatment.

## Methods

Nine different respirator models were tested (Table 1), most of which were certified as FFP3. Due to the lack of availability of FFP3 masks, some of the tested masks were FFP2 versions. One of each type of respirator was tested, with the exception of one model, which was tested by two different wearers. Each respirator was tested once by one of four wearers using the HSE INDG 479 (“Guidance on respiratory protective equipment (RPE) fit testing - INDG479,” 2019) and BSIF Fit2Fit Companion Guide for Quantitative Fit Test protocol (“Fit2Fit Companion to HSE INDG479,” n.d.) with a TSI PortaCount Pro+ 8038 (TSI Incorporated, Shoreview, MN USA), with a TSI Model 8026 Particle Generator which generates non-toxic NaCl particles. Three of the wearers were trained fit testers and male. One tester was female, and not a trained tester. The test protocol included 84 seconds of each of the following activities in order, conducted while walking in place: normal breathing; deep breathing; turning head side-to-side; nodding head up and down; reciting the rainbow passage; bending over (standing); and normal breathing. The fit factor (FF) is the ratio of ambient air particle concentration to in-respirator particle concentration and is calculated as:

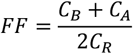

**Table 1:**
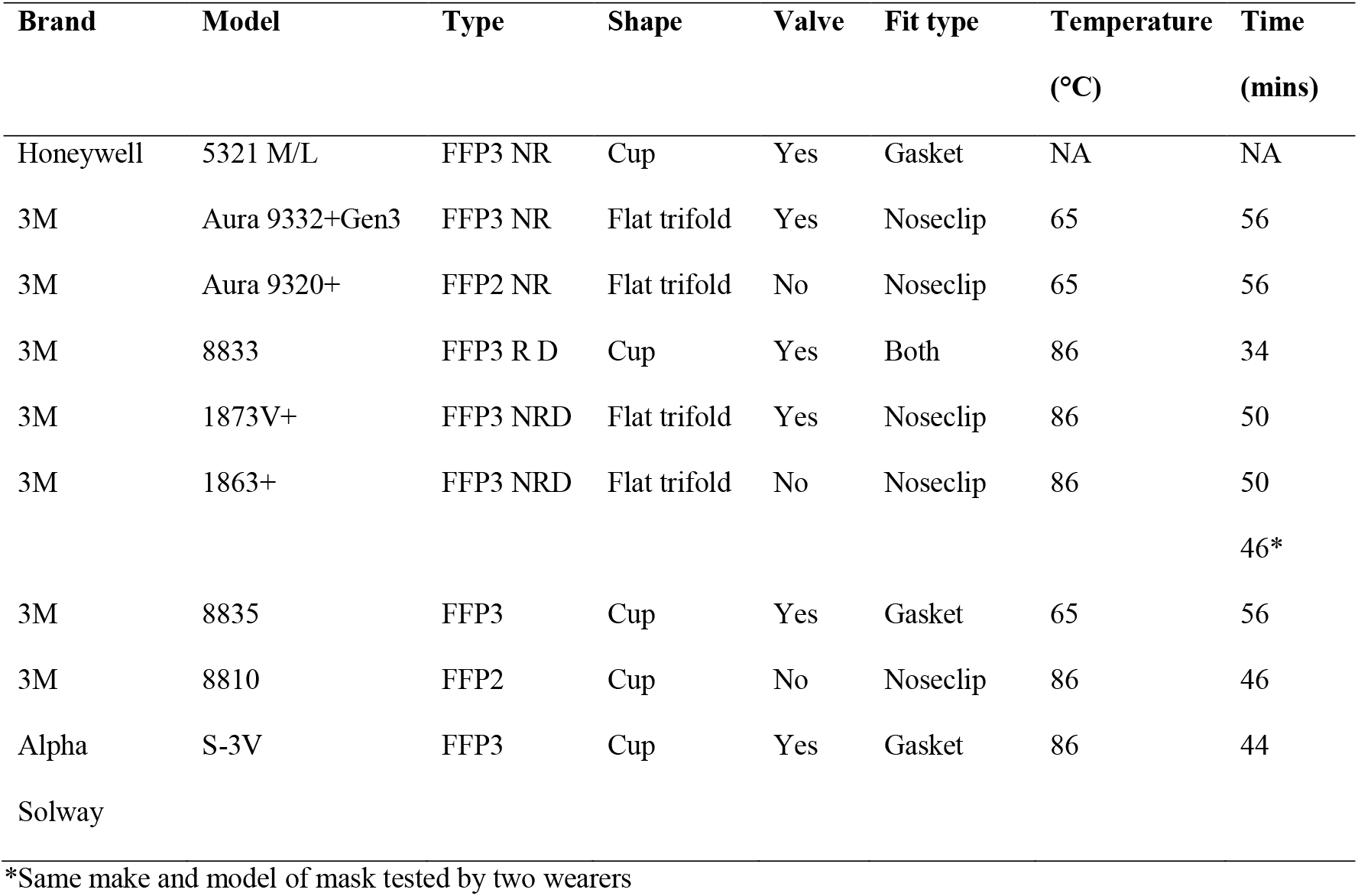
Masks tested, characteristics, oven temperature and time duration at that temperature

Where *C*_*B*_ = particle concentration in ambient air before respirator sample, *C*_*A*_ = particle concentration in ambient air after respirator sample, *C*_*R*_ = particle concentration in respirator.

The pass level was set at FF=100 and the test was stopped once failure was detected.

After the first fit test, respirators were then placed in a drying oven with a target of 85 degrees for approximately one hour, with a minimum of 30 minutes at the target temperature. Oven temperature was set using an external calibrated temperature probe. Respirators were placed in nylon heat resistant bags before placement in the oven. Two data logging temperature sensors (Hydrochron iButton® DS1923; Maxim Integrated, San Jose, CA, USA) were placed in the bags with the respirators to check the actual temperature in the vicinity of the respirator. The total time in the oven was generally about an hour due to time to equilibration for the temperature probes.

After removal from the oven, the respirators were allowed to cool for at least half an hour before re-test by the same wearer. Treatment generally occurred several hours to about a day after the initial test. Overall fit factor was compared pre- and post-treatment. It should be noted that experience with fit testing has shown that a degree of test-retest variability is to be expected and that this would be encompassed by the re-test results here. However, such variation would be expected to be random rather than systematic.

## Results

One respirator failed the initial pre-treatment fit test and was left out of the study. Two post-treatment tests were failed, while all others passed (Figure 1). With the exception of one respirator (3M 8810), all passing respirators had lower overall fit factors upon post-treatment tests. The reasonable consistency in the direction of change suggests this is a real effect and not an artefact created by random test-retest variability.

**Figure 1:**
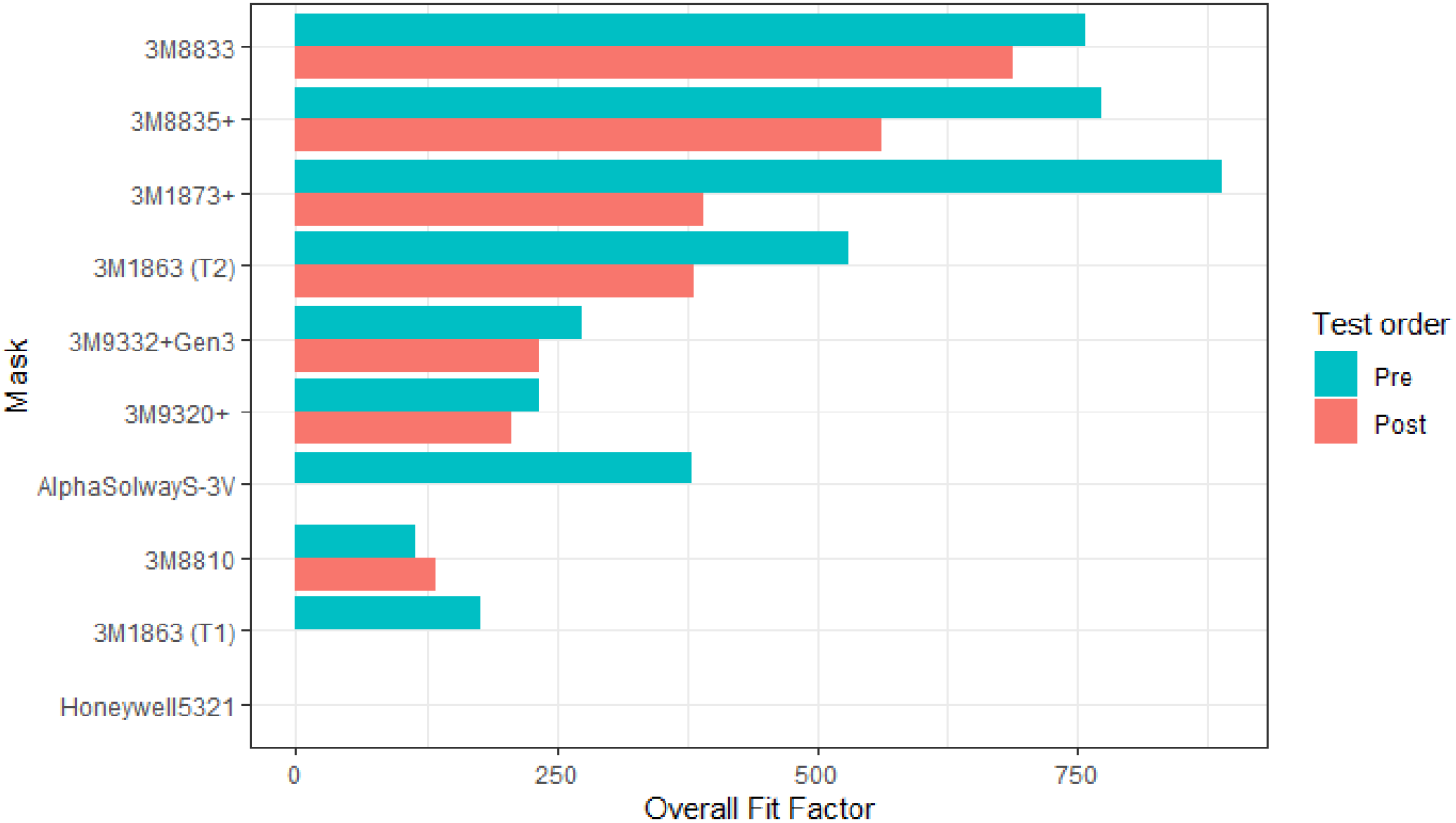
Overall fit factor of masks tested pre- and post- heat treatment (3M1863 was tested by two people)

During one of the heat treatments, one of the temperature loggers read a maximum temperature of 65*°C* even as the other logger in the oven at the same time, read 86°C. This is likely to have affected at least two respirators (Table 1).

## Discussion

We have examined the ability of FFP2/FFP3 respirators to pass a quantitative fit test after a single wear and heat treatment. The heat treatments were all at or above temperatures shown in other studies to deactivate the SARS-CoV-2 virus. Most respirators, of both flat fold and moulded cup style, were able to pass a re-test.

In this study, because we were testing donated respirators, most of our wearers were not previously fit tested for these respirators, with the exception of one. Although we did not test filtration efficiency post-heat treatment, it is unlikely that this was adversely affected by the treatment, as most respirators passed the second fit test. One of the respirator models that failed post-treatment fit test was duplicate tested by a different wearer, who passed the post-treatment test, therefore the fit test failure was likely due to alterations in fit, not filtration efficiency. In both failed fit tests post-treatment, visual inspection of the respirators indicated that some physical aspect of the respirator was adversely affected. For the 3M 1863 failure – the wearer had difficulty with refitting the nose clip the second time. For the Alpha Solway S-3V failure – the foam gasket around the face seal appeared to be more wrinkled and loose after the first wear. It is likely, therefore that impaired fit is responsible for both these failures, rather than filtration efficiency.

The post-treatment fit tests tended to have a lower overall fit factor, indicating that there may be some degradation, although this is not a large enough sample to determine whether this is a real effect or a chance finding. Repeated treatments and fit tests on the same respirator would need to be done to determine the likelihood that FFP respirators could be treated and re-used effectively multiple times. Our study shows that it is possible to treat and re-use FFP respirators at least once, thus doubling the potential numbers of respirator uses. This does not take into account any wear/deformation during use that might impair the extent to which it can be re-used. For example, these tests were conducted for a relatively short wear time, and in real use respirators may be worn for longer periods, and this may also affect the reusability of the mask.

Finally, although we were able to show that FFP respirators can withstand heat treatment and still pass a quantitative fit test, we did not test actual pathogen decontamination. Even if SARS-CoV-2 can be successfully decontaminated at <70°C, this does not mean that other pathogens or harmful microbes are effectively deactivated. Future work should include a pathogen viability test, and a greater range of temperatures. We were unable to measure humidity, and more controlled temperature/humidity combinations should be tested to optimise both decontamination and preservation of respirator effectiveness.

## Conclusions

Heat treatment at temperatures which inactivate the SARS-CoV-2 virus did not adversely affect filtration or fit of most FFP2 or FFP3 respirators tested, although we acknowledge that there is still a reasonable risk of fit failure. Heat could be a simple and inexpensive means to decontaminate respirators, and further investigation is needed to understand the impact of repeated long wear-decontamination-reuse cycles on respirator performance. Furthermore, we need to determine the optimal temperature and humidity conditions for acceptable decontamination while still retaining filtration and fit of the respirator. We strongly recommend that reuse of respirators, especially those which require more deformation to fit the user be kept with the original individual. Users should be taught how to visually inspect the respirators and how to do a seal check before re-use.

## Data Availability

Data are available upon request.

## Acknowledgements

We thank Tim Small, Alexis Dole, Calum Craig, and others who have donated FFP respirators for testing, and especially to Tim who approached us to do this. We also give a big thanks to Craig Lewis, and David Holmes, who did the face fit testing amidst their busy schedules. Thank you also to the IOM laboratory team who provided the oven facilities, especially to David Todd who helped facilitate this. Thanks to Karen Galea and Richard Gravelling for reviewing this work.

